# Extraction of Glaucoma Diagnosis, Type, and Severity from Clinical Notes using Secure Cloud-based Large Language Models

**DOI:** 10.64898/2026.06.18.26355532

**Authors:** Gustavo A. Samico, Nicholas Solages, Rafael Scherer, Rohit Muralidhar, Naomi E. Gutkind, Vitoria Palazoni, Felipe A. Medeiros, Swarup S. Swaminathan

## Abstract

**Purpose:** To evaluate the performance of large language models (LLMs) in extracting glaucoma diagnosis, type, and severity from clinical notes in the electronic health record (EHR).

**Methods:** Clinical notes of glaucoma-related encounters between 2014 and 2024 from 1,242 subjects were extracted from the Bascom Palmer Ophthalmic Repository. Two fellowship-trained glaucoma specialists labeled notes for glaucoma presence, type, and severity. Dataset was split into development (10%), validation (10%), and test (80%) sets. Five LLMs (Claude Opus 4.6, DeepSeek-V3.2, GPT-5.2, Grok 4.1 fast non-reasoning, and Qwen3.6-35B-A3B) were accessed via Microsoft Foundry within HIPAA-compliant containers. Model performance was assessed using standard metrics. Clinician-entered ICD-10 codes were also compared with adjudicated labels.

**Main Outcome Measures:** Gwet AC1, accuracy, sensitivity, specificity, and F1-score.

**Results:** Inter-grader agreement for glaucoma diagnosis, type, and severity was high (Gwet AC1 0.940, 0.917, 0.906, respectively). For glaucoma diagnosis, LLMs demonstrated high accuracy, with Claude achieving 97.5%, DeepSeek 96.0%, GPT 96.2%, Grok 94.4%, and Qwen 95.5%. F1 scores for glaucoma detection ranged from 97.0% to 98.9%. For type, accuracies were 97.1%, 94.2%, 94.2%, 94.0%, and 94.4%, respectively. F1 scores for the most prevalent type (POAG) ranged from 96.6% to 98.9%. For severity, accuracies were 95.0%, 94.8%, 94.5%, 94.0%, and 95.2%, respectively, with F1 scores ranging from 89.7% to 96.3%. ICD-10 codes demonstrated lower accuracies for type and severity (87.3% and 69.2%, respectively).

**Conclusions:** LLMs accurately extracted glaucoma diagnosis, type, and severity from free-text clinical notes, outperforming ICD-based phenotyping.

**Translational Relevance:** LLMs have the potential to transform unstructured documentation into scalable, research-ready phenotypes for EHR-based glaucoma cohort construction.

## INTRODUCTION

Glaucoma remains the leading cause of irreversible blindness worldwide, imposing a burden on aging populations.^1^ By 2040, more than 100 million people are projected to be affected, highlighting the importance of developing scalable strategies that ensure high accuracy and precision in clinical management as well as research to guide clinical care.^2^ Disease management depends on accurate characterization of glaucoma diagnosis, type, and severity, information that is frequently documented within electronic health records (EHRs), particularly in narrative clinical notes.^3, 4^

The widespread adoption of EHRs has enabled large-scale ophthalmic research, but most studies rely primarily on structured data elements such as International Classification of Diseases (ICD) codes.^4, 5^ Although convenient, ICD-based phenotyping can be inaccurate for glaucoma type and severity classification, potentially introducing bias into epidemiologic studies and cohort development.^6, 7^ These limitations are not merely administrative, as inaccurate phenotyping can distort prevalence estimates, compromise cohort validity, and ultimately impair the quality of evidence derived from large-scale EHR-based studies. Clinically relevant information often exists only within free-text documentation, limiting its accessibility for research.^8, 9^

Natural language processing (NLP) methods have been used to extract information from ophthalmology notes, including glaucoma diagnoses and medication data.^8, 10–12^ However, many traditional NLP approaches require task-specific development, extensive customization, or large annotated datasets, limiting scalability across clinical applications. Furthermore, traditional NLP systems generally depend on explicit documentation patterns or rule-based extraction methods, limiting their ability to interpret nuanced clinical reasoning embedded within narrative notes.^10, 12^ Prior ophthalmic NLP studies have primarily focused on binary disease identification or broad disease categorization but not on the extraction of disease severity, which is clinically meaningful in research cohorts and a key component of disease characterization.^11^ Consequently, scalable methods capable of generating detailed glaucoma phenotypes directly from routine clinical documentation remain limited.

Recent advances in large language models (LLMs) offer a complementary paradigm for clinical phenotyping from unstructured EHR data. Rather than requiring specialty-specific pretraining or customized extraction pipelines, contemporary LLMs can perform complex information extraction through prompt engineering alone, enabling rapid deployment across institutions and clinical applications.^13^ The purpose of this study was to evaluate whether secure cloud-based LLMs can accurately and comprehensively characterize glaucoma phenotypes by extracting glaucoma diagnosis, type, and disease severity from free-text clinical notes. We additionally compared LLM-derived phenotypes with clinician-entered ICD-10 classifications. We hypothesized that LLMs would demonstrate high concordance with expert clinician adjudication and outperform ICD-based phenotyping approaches. If confirmed, these findings would support the use of secure cloud-based LLMs as a practical and scalable strategy for improving glaucoma phenotyping in large-scale EHR-based research.

## METHODS

### Data Collection and Note Verification

This study was approved by the Institutional Review Board at the University of Miami. The requirement for informed consent was waived because of the retrospective nature of the study. The procedures and protocols followed during the study adhere to the Declaration of Helsinki and comply with the Health Insurance Portability and Accountability Act (HIPAA) for maintaining patient confidentiality and integrity.

This study was a retrospective analysis of patients from the Bascom Palmer Ophthalmic Repository (BPOR), which contains data from over 70,000 patients cared for at the Bascom Palmer Eye Institute (BPEI).^14, 15^ This database contains demographic, exam, imaging, procedural, and clinical note data of eyes with glaucoma or suspected of having glaucoma examined at BPEI, identified using ICD codes from the EHR system (Epic Systems, Verona, WI).

Prior work details the extraction and classification of clinical notes in BPOR.^16^ Briefly, we included medical notes recorded between 2014 and 2024 from outpatient encounters with finalized notes and glaucoma-related ICD codes (H40.X) listed in the visit diagnoses. Notes were stored on a HIPAA-compliant institutional Box server (Box, Redwood City, CA).

Notes were filtered to include only outpatient visit encounters with clinically relevant appointment types (e.g., new patient, follow-up, in-house referral visits). Notes corresponding to hybrid visits or preoperative encounters were excluded given their limited detail on disease staging or management plans. To ensure adequate data quality, we calculated text length for all notes and excluded those with fewer than 200 characters. After filtering, unique identifiers were used to prevent duplication across medical record numbers (MRNs), note IDs, and encounter IDs. The final curated dataset contained 1,250 notes, representing 1,250 unique patients, with one encounter and note per patient. After exclusion of eight physician attestations that lacked independent clinical content, 1,242 notes remained available for annotation and model evaluation. Note lengths ranged from 208 to 7,633 characters, with a mean of approximately 2,046 characters. Notes were stratified by clinician to ensure a heterogeneous mixture of writing styles.

We extracted all clinician-entered International Classification of Diseases, 10th Revision, Clinical Modification (ICD-10-CM) codes for visit diagnoses beginning with H40.X, as entered by clinicians in the EHR. ICD codes were systematically parsed to separate glaucoma diagnosis, type, and severity for each eye. When laterality was unspecified (codes containing "X" placeholders or digits "0" or "9"), cases were considered bilateral and assigned to both eyes. When evaluating glaucoma type, clinician-entered ICD-10 codes were grouped into the following phenotypes: "Primary Open-Angle Glaucoma" (POAG; H40.11X), "Normal Tension Glaucoma" (NTG; H40.12X), "Pigmentary Glaucoma" (PG; H40.13X), "Pseudoexfoliative Glaucoma" (PXG; H40.14X), "Primary Angle-Closure Glaucoma" (PACG; H40.2X), "Angle Recession Glaucoma" (H40.3X), "Uveitic Glaucoma" (H40.4X), and "Glaucoma Suspect" (H40.0X). Because of the absence of unique ICD categories for some glaucoma types (e.g., mixed mechanism glaucoma, neovascular glaucoma), ICD-level performance metrics were calculated only for diagnoses with specific and unambiguous ICD mappings as noted.

### Note Labeling

All notes were independently reviewed by two fellowship-trained glaucoma specialists for glaucoma diagnosis, type, and severity at the eye level. In cases of disagreement between the two graders, a third glaucoma specialist served as an adjudicator to establish the final reference standard. This process established a "ground truth" benchmark for each extraction task, which was subsequently used to evaluate model performance. For each encounter note, labelers classified each eye as “glaucoma”, “glaucoma suspect”, or “neither”. Disease type was labeled as one of the following categories: POAG, PACG, PXG, PG, NTG, Steroid, Uveitic, Mixed Mechanism, Angle Recession, Neovascular, Congenital, Sturge–Weber, Glaucoma following cataract surgery, Iridocorneal endothelial (ICE) syndrome, Unspecified, Other, or N/A. Glaucoma suspects were categorized as “N/A”. If two types were listed, the non-POAG or non-PACG type was used as the label. If two types were listed with one being noted just as a possible “component”, the main type was used as the label. If type could not be ascertained or there was significant contradiction within the note, graders were instructed to label as “Unspecified”. Severity was recorded as Mild, Moderate, Severe, or Other. Labeling was based on the mention of severity stage by the physician.^17^ If there was a mention of preperimetric disease, it was defined as mild. If the note used the terms “end-stage” or “advanced stage”, that eye was classified as severe. If more than one stage were mentioned, the more advanced stage was used as the label. Given the goal of conservative phenotyping, we required explicit clinician confirmation of the stage (i.e., mention of structural or perimetric findings without an explicit stage descriptor was not used). If severity could not be ascertained even after evaluating for these conditions or the eye were noted to be a suspect or non-glaucomatous, graders were instructed to label as “Other”. When laterality was not explicitly specified in the clinical note, the diagnosis was conservatively assigned to both eyes as previously completed and as was done with ICD-10 codes as noted above.^18^

### Prompt Engineering

In contrast to traditional machine learning workflows, LLM parameters were not fine-tuned. Instead, the development set was only used for prompt engineering and choosing few-shot examples, which required only a small dataset. No model fine-tuning or parameter optimization was performed. The validation set was utilized to refine prompts further, after which prompts were frozen. The held-out test set was then used for analysis of LLM performance. This approach is consistent with prior work in which model behavior is optimized through iterative prompt-refinement rather than through parameter tuning.^19–21^ Prompts incorporated rule-based instructions, few-shot examples, checklists for verification, and structured reasoning decomposition inspired by chain-of-thought prompt engineering. The full text of the three prompts is available as **Supplementary Material.**

As described in the prompts, predefined rules were used to address potentially conflicting clinical documentation. Briefly, for glaucoma diagnosis, an explicitly documented diagnosis of glaucoma took precedence over glaucoma suspect status for the same eye. For glaucoma type, the primary diagnosis was prioritized when an additional type was described only as a suspected or contributing component. When conflicting types appeared in different sections of the note, the "Problem List Items Addressed This Visit" section was prioritized over the history of present illness (HPI).

When multiple glaucoma types were documented without a clear primary diagnosis, non-POAG and non-PACG diagnoses were prioritized, with unresolved contradictions classified as "Unspecified." For severity staging, when multiple stages were documented for the same eye, the more advanced stage was assigned. Cases in which severity could not be established according to the predefined criteria were classified as "Other." These decision rules were incorporated into the prompts.

The 1,242 clinical notes were divided into development (n = 125), validation (n = 125), and held-out test (n = 992) sets, stratified by clinician, corresponding to approximately 10%, 10%, and 80% of the dataset, respectively. The development set was used to guide prompt creation, with the validation set used to further refine the prompts. The held-out test set was used for assessment of model performance.

### Model Architecture and Setup

LLMs were accessed through a secure cloud-based infrastructure using Microsoft Foundry (Microsoft, Redmond, WA, USA). All assessments were conducted within HIPAA-compliant containers configured to protect protected health information (PHI) under a business associate agreement between Microsoft and the University of Miami. Notes remained within the secure cloud environment and were not transmitted to external services outside the protected infrastructure. From the LLMs available within the platform, we selected five models for evaluation: Claude Opus 4.6 (Anthropic, San Francisco, CA, USA), DeepSeek-V3.2 (DeepSeek AI, Hangzhou, China), GPT-5.2 (OpenAI, San Francisco, CA, USA), Grok 4.1 fast non-reasoning (xAI, Palo Alto, CA, USA), and Qwen3.6-35B-A3B (Alibaba Cloud, Hangzhou, China). Claude Opus 4.6 was accessed through an Anthropic Foundry interface, GPT-5.2 was accessed through the Azure OpenAI service, and DeepSeek, Grok, and Qwen were accessed through OpenAI-compatible APIs using model-specific endpoints. For each model, configuration parameters included deployment endpoint, model identifier, provider type, and available reasoning settings. To ensure standardized comparisons across models and minimize variability related to reasoning configurations, all models received identical prompts, a temperature of 0.4, top_p of 0.9, and maximum output length of 500 tokens. Qwen 3.6-35B-A3B was additionally configured with a top_k of 20. Because each model varied in its reasoning controls, reasoning was disabled using each model’s own mechanism.

Reasoning effort was set to “none” for GPT-5.2, while extended thinking was disabled for Claude Opus 4.6. Thinking was turned off for Qwen 3.6-35B-A3B by setting “enable_thinking” to false. Grok 4.1 Fast Non-Reasoning had no reasoning mode by design. DeepSeek-V3.2 was the only exception, because its lowest available reasoning setting was ‘minimal’ rather than ‘none’. Each clinical note was submitted with its corresponding encounter date and the task-specific prompt. Model outputs were then exported into task- and model-specific result files for downstream analysis.

Outputs were compared against expert-adjudicated reference labels. Model outputs were programmatically parsed into structured JavaScript Object Notation (JSON) format, with validation and error handling for improperly formatted responses. If a model’s response could not be parsed or contained no classification for either eye, the output was recorded as a failure as per prior studies. ^22, 23^ Both eyes from that note were excluded from further performance calculations and were reported as failure rates. No failures occurred due to missing labels. Predictions of categories that did not appear among the reference labels (e.g., congenital glaucoma) were retained and counted as incorrect. Reasoning for model outputs was also extracted as a parameter to provide insight into LLM analysis.

### Fine-Tuned Encoder Comparator

To compare prompt-based extraction with natural language processing (NLP), we fine-tuned BioClinicalBERT, a clinical encoder pretrained on medical notes,^24^ separately for each task. To match the annotated data available to the LLMs, fine-tuning used only the 250 notes from the development and validation sets, and evaluation used the held-out 992-note test set. Because labels were assigned per eye, each note was converted into two examples with identical text and an eye-identifying prefix (right or left eye). BioClinicalBERT accepts at most 512 tokens, which would have truncated 54.7% of notes before the assessment and plan, so notes were encoded with overlapping 512-token windows (256-token stride) combined by attention-weighted pooling. Class imbalance was addressed with an inverse-frequency class-weighted cross-entropy loss. All tasks used identical hyperparameters: AdamW optimizer, learning rate 4 × 10⁻ (selected by sweep), weight decay 0.01, warmup over the first 10% of steps, gradient clipping at norm 1.0, and effective batch size 16 (2 per step × 8 gradient-accumulation steps). Hyperparameters were evaluated with 5-fold cross-validation stratified by label and grouped by note, keeping both eyes of a subject in the same fold. Training ran for up to 15 epochs, stopping early after 3 evaluation rounds without improvement in validation macro-averaged F1 score. The final model for each task was refit on a random 90% of the 250 notes, with the remaining 10% (split by note) held out to monitor early stopping. Performance was evaluated at the eye level with the held-out test set using standard multiclass metrics (e.g., accuracy, balanced accuracy, macro F1, weighted F1, Cohen’s κ), with Wilson 95% confidence intervals (CIs) for class-specific sensitivity, specificity, PPV, and NPV and percentile bootstrap CIs for all other metrics.

### Statistical Analysis

Inter-grader agreement was assessed before adjudication using Gwet’s AC1, particularly useful when class imbalance is expected. Corresponding 95% CIs for glaucoma diagnosis, type classification, and severity staging were calculated. Agreement was calculated using the held-out test set.

All LLM performance analyses were conducted at the eye level. Model outputs of the test set were benchmarked against adjudicated labels, with assessments completed at the eye level. Performance metrics for each model were calculated using only eyes with valid model outputs (i.e., failures were excluded). For each LLM, we evaluated their metrics of each task – accuracy, sensitivity (recall), specificity, positive predictive value (PPV), negative predictive value (NPV), and F1 score were computed using adjudicated labels as the reference standard. For glaucoma diagnosis, multiclass performance was evaluated across the categories of glaucoma, suspect, and neither. For glaucoma type and severity classification, one-vs-rest analyses were performed for each group. For proportion-based performance metrics, including accuracy, sensitivity, specificity, positive predictive value, and negative predictive value, 95% CIs were calculated using the Wilson score method. CIs for F1 scores were estimated using bootstrap resampling for LLM-based classifications and approximated using the Wilson score method for ICD-10-CM-based classifications. ICD-10-derived classifications were evaluated using the same reference standard and performance metrics to allow direct comparison with LLM outputs. All statistical analyses and performance metrics were computed in Python (Python Software Foundation, Wilmington, DE, USA) using the scikit-learn, statsmodels, and pandas libraries.

## RESULTS

When evaluating the total dataset (1,242 notes or 2,484 eye-level labels), adjudication was required for 119 (4.8%) glaucoma diagnosis labels, 219 (8.8%) glaucoma type labels, and 283 (11.4%) severity labels. The held-out test set was comprised of 992 clinical notes (1,984 eye-level labels). Gwet’s AC1 was 0.940 (95% CI: 0.928–0.952) for diagnosis, 0.917 (95% CI: 0.904–0.930) for type, and 0.906 (95% CI: 0.891–0.921) for disease severity, with raw percent agreement of 95.4%, 92.2%, and 92.5%, respectively. A total of 69.4% of eyes were noted to have glaucoma, 24.1% suspected of glaucoma, and 6.5% neither. POAG was the most common glaucoma type, accounting for 800 eyes (58.4% of glaucomatous eyes; Table 1). Regarding disease severity, 15.7%, 9.2%, 18.0%, and 57.1% were noted to be mild, moderate, severe, and other, respectively. When excluding eyes identified as “suspect” or “neither” per adjudicated labels, 22.7%, 13.2%, 25.9%, and 38.2% were mild, moderate, severe, and other, respectively.

**Table 1.** Glaucoma type distribution by eye within the test set.

| Glaucoma Type | N | % of Total<br>(n = 1,984) | % of Classified<br>(n = 1,369) |
|---|---|---|---|
| POAG | 800 | 40.3 | 58.4 |
| NTG | 127 | 6.4 | 9.3 |
| PACG | 105 | 5.3 | 7.7 |
| Pseudoexfoliative Glaucoma | 97 | 4.9 | 7.1 |
| Mixed Mechanism Glaucoma | 66 | 3.3 | 4.8 |
| Unspecified Glaucoma | 44 | 2.2 | 3.2 |
| Neovascular Glaucoma | 31 | 1.6 | 2.3 |
| Other | 22 | 1.1 | 1.6 |
| Uveitic Glaucoma | 21 | 1.1 | 1.5 |
| Pigmentary Glaucoma | 20 | 1.0 | 1.5 |
| Steroid-induced Glaucoma | 15 | 0.8 | 1.1 |
| Angle Recession Glaucoma | 8 | 0.4 | 0.6 |
| Glaucoma following cataract surgery | 7 | 0.4 | 0.5 |
| ICE Syndrome | 5 | 0.3 | 0.4 |
| Sturge-Weber Glaucoma | 1 | 0.1 | 0.1 |
| Not classified (Suspect/Neither) | 615 | 31.0 | — |

Classified eyes include only those assigned a glaucoma type. ICE = Iridocorneal Endothelial; NTG = Normal Tension Glaucoma; PACG = Primary Angle-Closure Glaucoma; POAG = Primary Open-Angle Glaucoma.

For glaucoma diagnosis, overall accuracy ranged from 94.4% (95% CI: 93.3% - 95.3%) to 97.5% (95% CI: 96.7% - 98.1%) across the five evaluated models. Claude achieved the highest performance (97.5%; 95% CI: 96.7% - 98.1%), followed by GPT (96.2%; 95% CI: 95.3% - 97.0%), DeepSeek (96.0%; 95% CI: 95.0% - 96.7%), Qwen (95.5%; 95% CI: 94.5% - 96.3%), and Grok (94.4%; 95% CI: 93.3% - 95.3%). Failure rates ranged from 0% to 2.0% of notes across models, with Claude and Qwen having the most failures. **Figure 1** illustrates the confusion matrix using Claude outputs. Class-specific performance metrics are shown in **Table 2**. ICD-10 codes had a 92.0% sensitivity and 77.3% specificity for glaucoma detection, with an accuracy of 87.5% (95% CI: 86.0% - 88.9%) and F1 score of 91.1% (95% CI: 90.0% - 92.1%). Glaucoma suspect identification with ICD-10 codes had a 70.2% sensitivity and 96.5% specificity, with an accuracy of 90.1% (95% CI: 88.7% - 91.4%) and F1 score of 77.4% (95% CI: 74.5% - 80.1%).

**Figure 1.**
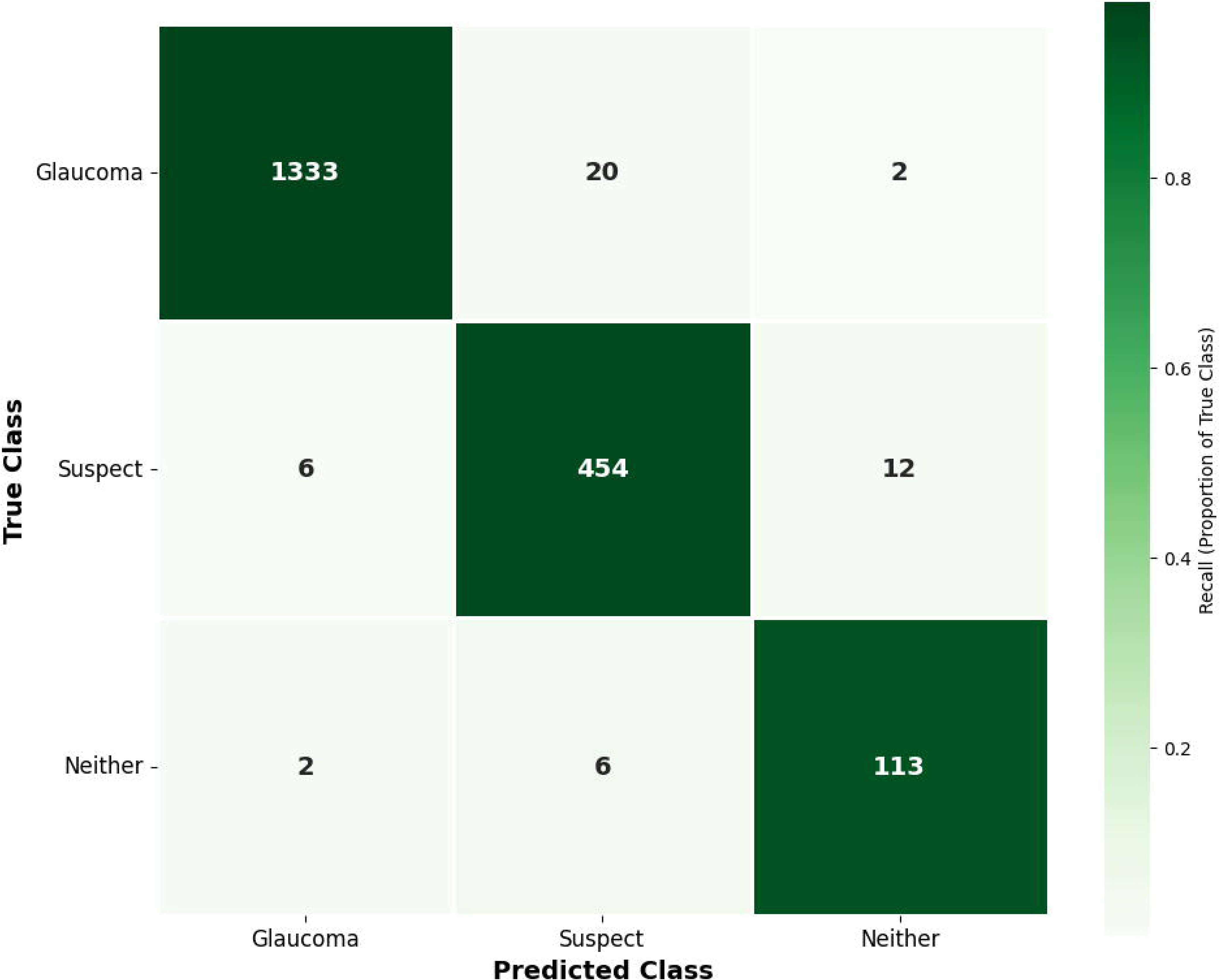
Confusion matrix for glaucoma diagnosis using output from Claude. A total of 36 eyes from 18 notes with failed outputs were excluded.

**Table 2.**
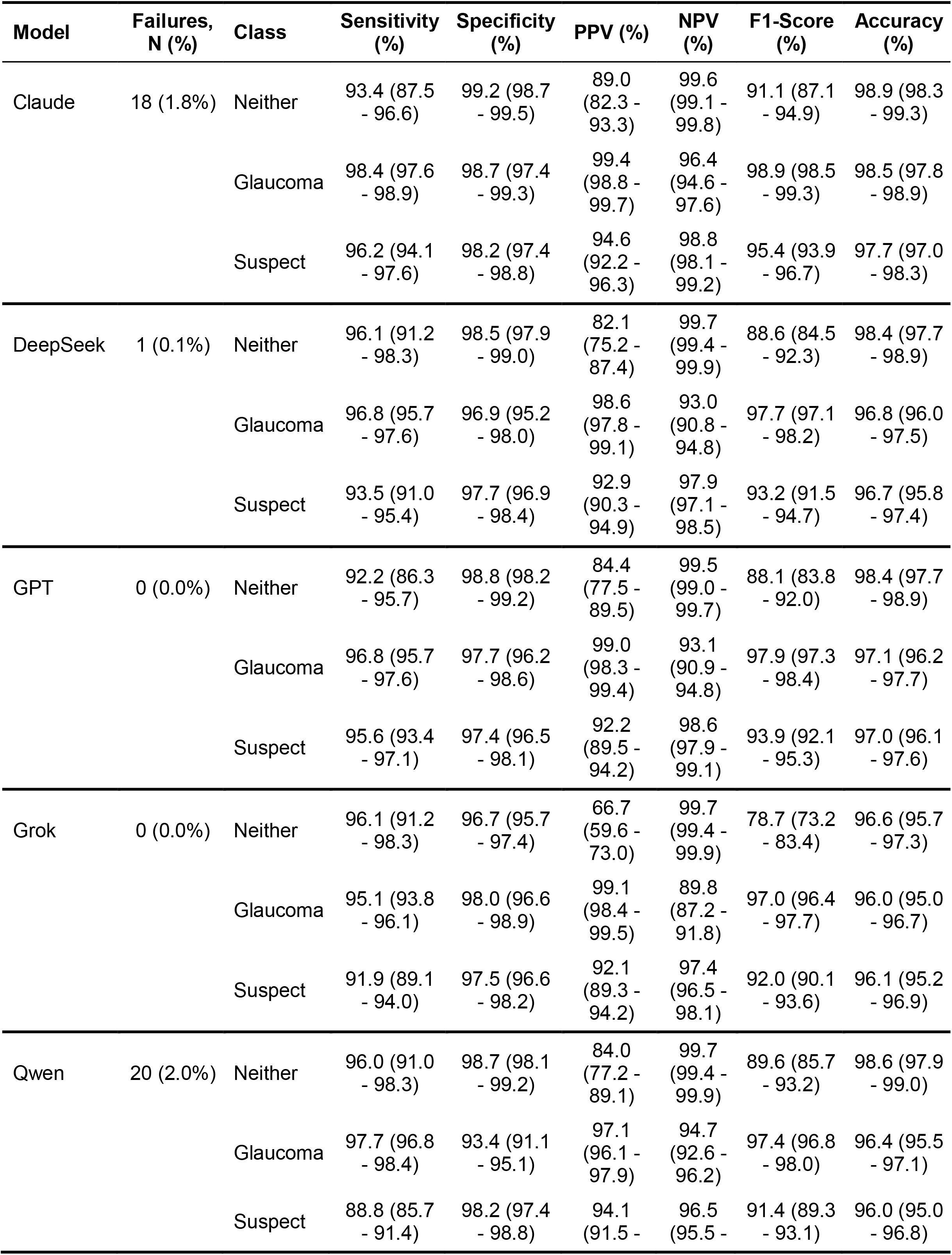
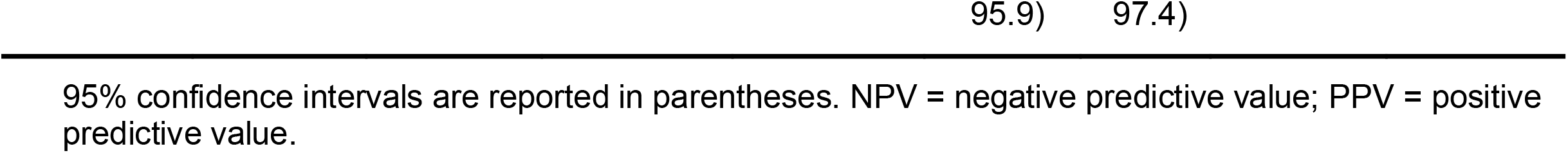
Class-specific performance of large language models for glaucoma diagnosis.

| Model | Failures, N (%) | Class | Sensitivity (%) | Specificity (%) | PPV (%) | NPV (%) | F1-Score (%) | Accuracy (%) |
|---|---|---|---|---|---|---|---|---|
| Claude | 18 (1.8%) | Neither | 93.4 (87.5 - 96.6) | 99.2 (98.7 - 99.5) | 89.0 (82.3 - 93.3) | 99.6 (99.1 - 99.8) | 91.1 (87.1 - 94.9) | 98.9 (98.3 - 99.3) |
|  |  | Glaucoma | 98.4 (97.6 - 98.9) | 98.7 (97.4 - 99.3) | 99.4 (98.8 - 99.7) | 96.4 (94.6 - 97.6) | 98.9 (98.5 - 99.3) | 98.5 (97.8 - 98.9) |
|  |  | Suspect | 96.2 (94.1 - 97.6) | 98.2 (97.4 - 98.8) | 94.6 (92.2 - 96.3) | 98.8 (98.1 - 99.2) | 95.4 (93.9 - 96.7) | 97.7 (97.0 - 98.3) |
| DeepSeek | 1 (0.1%) | Neither | 96.1 (91.2 - 98.3) | 98.5 (97.9 - 99.0) | 82.1 (75.2 - 87.4) | 99.7 (99.4 - 99.9) | 88.6 (84.5 - 92.3) | 98.4 (97.7 - 98.9) |
|  |  | Glaucoma | 96.8 (95.7 - 97.6) | 96.9 (95.2 - 98.0) | 98.6 (97.8 - 99.1) | 93.0 (90.8 - 94.8) | 97.7 (97.1 - 98.2) | 96.8 (96.0 - 97.5) |
|  |  | Suspect | 93.5 (91.0 - 95.4) | 97.7 (96.9 - 98.4) | 92.9 (90.3 - 94.9) | 97.9 (97.1 - 98.5) | 93.2 (91.5 - 94.7) | 96.7 (95.8 - 97.4) |
| GPT | 0 (0.0%) | Neither | 92.2 (86.3 - 95.7) | 98.8 (98.2 - 99.2) | 84.4 (77.5 - 89.5) | 99.5 (99.0 - 99.7) | 88.1 (83.8 - 92.0) | 98.4 (97.7 - 98.9) |
|  |  | Glaucoma | 96.8 (95.7 - 97.6) | 97.7 (96.2 - 98.6) | 99.0 (98.3 - 99.4) | 93.1 (90.9 - 94.8) | 97.9 (97.3 - 98.4) | 97.1 (96.2 - 97.7) |
|  |  | Suspect | 95.6 (93.4 - 97.1) | 97.4 (96.5 - 98.1) | 92.2 (89.5 - 94.2) | 98.6 (97.9 - 99.1) | 93.9 (92.1 - 95.3) | 97.0 (96.1 - 97.6) |
| Grok | 0 (0.0%) | Neither | 96.1 (91.2 - 98.3) | 96.7 (95.7 - 97.4) | 66.7 (59.6 - 73.0) | 99.7 (99.4 - 99.9) | 78.7 (73.2 - 83.4) | 96.6 (95.7 - 97.3) |
|  |  | Glaucoma | 95.1 (93.8 - 96.1) | 98.0 (96.6 - 98.9) | 99.1 (98.4 - 99.5) | 89.8 (87.2 - 91.8) | 97.0 (96.4 - 97.7) | 96.0 (95.0 - 96.7) |
|  |  | Suspect | 91.9 (89.1 - 94.0) | 97.5 (96.6 - 98.2) | 92.1 (89.3 - 94.2) | 97.4 (96.5 - 98.1) | 92.0 (90.1 - 93.6) | 96.1 (95.2 - 96.9) |
| Qwen | 20 (2.0%) | Neither | 96.0 (91.0 - 98.3) | 98.7 (98.1 - 99.2) | 84.0 (77.2 - 89.1) | 99.7 (99.4 - 99.9) | 89.6 (85.7 - 93.2) | 98.6 (97.9 - 99.0) |
|  |  | Glaucoma | 97.7 (96.8 - 98.4) | 93.4 (91.1 - 95.1) | 97.1 (96.1 - 97.9) | 94.7 (92.6 - 96.2) | 97.4 (96.8 - 98.0) | 96.4 (95.5 - 97.1) |
|  |  | Suspect | 88.8 (85.7 - 91.4) | 98.2 (97.4 - 98.8) | 94.1 (91.5 - 95.9) | 96.5 (95.5 - 97.4) | 91.4 (89.3 - 93.1) | 96.0 (95.0 - 96.8) |

95% confidence intervals are reported in parentheses. NPV = negative predictive value; PPV = positive predictive value.

For glaucoma type classification, overall accuracy ranged from 94.0% (95% CI: 92.9% - 95.0%) to 97.1% (95% CI: 96.3% - 97.8%). Individual model accuracies were 97.1% (95% CI: 96.3% - 97.8%), 94.2% (95% CI: 93.1% - 95.2%), 94.2% (95% CI: 93.0% - 95.1%), 94.0% (95% CI: 92.9% - 95.0%), and 94.4% (95% CI: 93.3% - 95.4%) for Claude, DeepSeek, GPT, Grok and Qwen, respectively. Failure rates remained low, ranging from 0% (GPT, Grok) to 5.2% (Qwen) across models. **Figure 2** presents the confusion matrix for glaucoma type classification using Claude outputs. Detailed class-specific performance metrics are shown in **Table 3**. When comparing ICD-10 diagnostic codes against adjudicated labels, overall accuracy was 87.3% (95% CI 85.2 - 89.0%) and macro-F1 was 77.7% (95% CI 71.9 – 82.0%). Diagnostic performance of ICD codes varied across glaucoma types as shown in **Table 4**. F1 scores ranged from 63.8% (uveitic glaucoma) to 88.1% (POAG).

**Figure 2.**
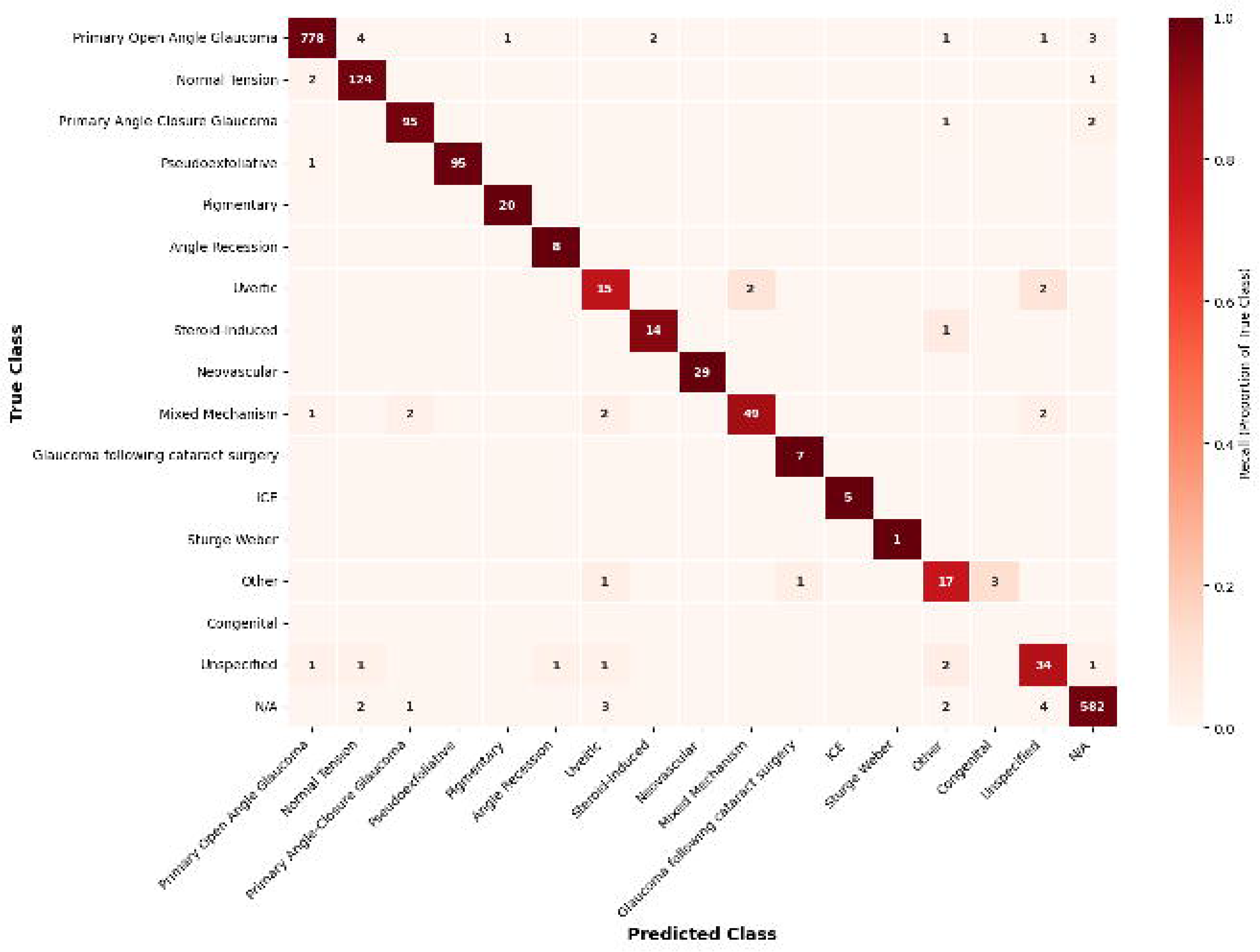
Confusion matrix for glaucoma type classification using output from Claude. A total of 56 eyes from 28 notes with failed outputs were excluded.

**Table 3.** Class-specific performance of large language models for glaucoma type classification.

| **Model** | **Failures, N (%)** | **Type** | **Sensitivity (%)** | **Specificity (%)** | **PPV (%)** | **NPV (%)** | **F1-Score (%)** | **Accuracy (%)** |
|---|---|---|---|---|---|---|---|---|
| Claude | 28 (2.8%) | POAG | 98.5 (97.4 - 99.1) | 99.6 (99.0 - 99.8) | 99.4 (98.5 - 99.7) | 99.0 (98.2 - 99.4) | 98.9 (98.4 - 99.4) | 99.1 (98.6 - 99.4) |
|  |  | NTG | 97.6 (93.3 - 99.2) | 99.6 (99.2 - 99.8) | 94.7 (89.4 - 97.4) | 99.8 (99.5 - 99.9) | 96.1 (93.6 - 98.3) | 99.5 (99.0 - 99.7) |
|  |  | PACG | 96.9 (91.4 - 99.0) | 99.8 (99.5 - 99.9) | 96.9 (91.4 - 99.0) | 99.8 (99.5 - 99.9) | 96.9 (94.1 - 99.1) | 99.7 (99.3 - 99.9) |
|  |  | Pseudoexfoliative Glaucoma | 99.0 (94.3 - 99.8) | 100.0 (99.8 - 100.0) | 100.0 (96.1 - 100.0) | 99.9 (99.7 - 100.0) | 99.5 (98.3 - 100.0) | 99.9 (99.7 - 100.0) |
|  |  | Pigmentary Glaucoma | 100.0 (83.9 - 100.0) | 99.9 (99.7 - 100.0) | 95.2 (77.3 - 99.2) | 100.0 (99.8 - 100.0) | 97.6 (91.4 - 100.0) | 99.9 (99.7 - 100.0) |
|  |  | Angle Recession Glaucoma | 100.0 (67.6 - 100.0) | 99.9 (99.7 - 100.0) | 88.9 (56.5 - 98.0) | 100.0 (99.8 - 100.0) | 94.1 (78.2 - 100.0) | 99.9 (99.7 - 100.0) |
|  |  | Uveitic Glaucoma | 78.9 (56.7 - 91.5) | 99.6 (99.2 - 99.8) | 68.2 (47.3 - 83.6) | 99.8 (99.5 - 99.9) | 73.2 (57.1 - 85.7) | 99.4 (99.0 - 99.7) |
|  |  | Steroid-Induced Glaucoma | 93.3 (70.2 - 98.8) | 99.9 (99.6 - 100.0) | 87.5 (64.0 - 96.5) | 99.9 (99.7 - 100.0) | 90.3 (76.9 - 100.0) | 99.8 (99.5 - 99.9) |
|  |  | Neovascular Glaucoma | 100.0 (88.3 - 100.0) | 100.0 (99.8 - 100.0) | 100.0 (88.3 - 100.0) | 100.0 (99.8 - 100.0) | 100.0 (100.0 - 100.0) | 100.0 (99.8 - 100.0) |
|  |  | Mixed Mechanism Glaucoma | 87.5 (76.4 - 93.8) | 99.9 (99.6 - 100.0) | 96.1 (86.8 - 98.9) | 99.6 (99.2 - 99.8) | 91.6 (85.7 - 96.4) | 99.5 (99.1 - 99.8) |
|  |  | Glaucoma following cataract surgery | 100.0 (64.6 - 100.0) | 99.9 (99.7 - 100.0) | 87.5 (52.9 - 97.8) | 100.0 (99.8 - 100.0) | 93.3 (72.7 - 100.0) | 99.9 (99.7 - 100.0) |
|  |  | ICE Syndrome | 100.0 (56.6 - 100.0) | 100.0 (99.8 - 100.0) | 100.0 (56.6 - 100.0) | 100.0 (99.8 - 100.0) | 100.0 (100.0 - 100.0) | 100.0 (99.8 - 100.0) |
|  |  | Sturge-Weber Glaucoma | 100.0 (20.7 - 100.0) | 100.0 (99.8 - 100.0) | 100.0 (20.7 - 100.0) | 100.0 (99.8 - 100.0) | 100.0 (100.0 - 100.0) | 100.0 (99.8 - 100.0) |
|  |  | Other | 77.3 (56.6 - 89.9) | 99.6 (99.2 - 99.8) | 70.8 (50.8 - 85.1) | 99.7 (99.4 - 99.9) | 73.9 (56.2 - 86.8) | 99.4 (98.9 - 99.6) |
|  |  | Congenital Glaucoma | — | 99.8 (99.5 - 99.9) | 0.0 (0.0 - 56.1) | 100.0 (99.8 - 100.0) | — | 99.8 (99.5 - 99.9) |
|  |  | Unspecified Glaucoma | 82.9 (68.7 - 91.5) | 99.5 (99.1 - 99.7) | 79.1 (64.8 - 88.6) | 99.6 (99.2 - 99.8) | 81.0 (71.0 - 89.4) | 99.2 (98.7 - 99.5) |
|  |  | N/A | 98.0 (96.5 - 98.8) | 99.5 (98.9 - 99.7) | 98.8 (97.6 - 99.4) | 99.1 (98.4 - 99.5) | 98.4 (97.6 - 99.1) | 99.0 (98.5 - 99.4) |
| DeepSeek | 2 (0.2%) | POAG | 97.6 (96.3 - 98.5) | 97.5 (96.4 - 98.2) | 96.3 (94.8 - 97.4) | 98.4 (97.5 - 99.0) | 97.0 (96.1 - 97.8) | 97.5 (96.7 - 98.1) |
|  |  | NTG | 96.1 (91.1 - 98.3) | 99.3 (98.8 - 99.6) | 90.4 (84.2 - 94.3) | 99.7 (99.4 - 99.9) | 93.1 (89.6 - 96.0) | 99.1 (98.6 - 99.4) |
|  |  | PACG | 94.3 (88.1 - 97.4) | 99.5 (99.1 - 99.7) | 91.7 (84.9 - 95.6) | 99.7 (99.3 - 99.9) | 93.0 (89.2 - 96.2) | 99.2 (98.8 - 99.5) |
|  |  | Pseudoexfoliative Glaucoma | 89.6 (81.9 - 94.2) | 99.9 (99.7 - 100.0) | 98.9 (93.8 - 99.8) | 99.5 (99.0 - 99.7) | 94.0 (90.2 - 97.2) | 99.4 (99.0 - 99.7) |
|  |  | Pigmentary Glaucoma | 100.0 (83.9 - 100.0) | 100.0 (99.8 - 100.0) | 100.0 (83.9 - 100.0) | 100.0 (99.8 - 100.0) | 100.0 (100.0 - 100.0) | 100.0 (99.8 - 100.0) |
|  |  | Angle Recession Glaucoma | 87.5 (52.9 - 97.8) | 99.9 (99.7 - 100.0) | 87.5 (52.9 - 97.8) | 99.9 (99.7 - 100.0) | 87.5 (66.7 - 100.0) | 99.9 (99.6 - 100.0) |
|  |  | Uveitic Glaucoma | 90.5 (71.1 - 97.3) | 99.8 (99.6 - 99.9) | 86.4 (66.7 - 95.3) | 99.9 (99.6 - 100.0) | 88.4 (76.5 - 96.9) | 99.7 (99.4 - 99.9) |
|  |  | Steroid-Induced Glaucoma | 61.5 (35.5 - 82.3) | 99.9 (99.6 - 100.0) | 80.0 (49.0 - 94.3) | 99.7 (99.4 - 99.9) | 69.6 (42.1 - 88.2) | 99.6 (99.3 - 99.8) |
|  |  | Neovascular Glaucoma | 96.8 (83.8 - 99.4) | 100.0 (99.8 - 100.0) | 100.0 (88.6 - 100.0) | 99.9 (99.7 - 100.0) | 98.4 (93.7 - 100.0) | 99.9 (99.7 - 100.0) |
|  |  | Mixed Mechanism Glaucoma | 83.1 (72.2 - 90.3) | 99.9 (99.7 - 100.0) | 98.2 (90.4 - 99.7) | 99.4 (99.0 - 99.7) | 90.0 (84.4 - 94.8) | 99.4 (98.9 - 99.7) |
|  |  | Glaucoma following cataract surgery | 100.0 (64.6 - 100.0) | 100.0 (99.8 - 100.0) | 100.0 (64.6 - 100.0) | 100.0 (99.8 - 100.0) | 100.0 (100.0 - 100.0) | 100.0 (99.8 - 100.0) |
|  |  | ICE Syndrome | 60.0 (23.1 - 88.2) | 100.0 (99.8 - 100.0) | 100.0 (43.9 - 100.0) | 99.9 (99.6 - 100.0) | 75.0 (0.0 - 100.0) | 99.9 (99.6 - 100.0) |
|  |  | Sturge-Weber Glaucoma | 100.0 (20.7 - 100.0) | 100.0 (99.8 - 100.0) | 100.0 (20.7 - 100.0) | 100.0 (99.8 - 100.0) | 100.0 (100.0 - 100.0) | 100.0 (99.8 - 100.0) |
|  |  | Other | 90.9 (72.2 - 97.5) | 99.1 (98.6 - 99.4) | 52.6 (37.3 - 67.5) | 99.9 (99.6 - 100.0) | 66.7 (50.8 - 79.4) | 99.0 (98.4 - 99.3) |
|  |  | Congenital Glaucoma | — | 99.9 (99.7 - 100.0) | 0.0 (0.0 - 79.3) | 100.0 (99.8 - 100.0) | — | 99.9 (99.7 - 100.0) |
|  |  | Unspecified Glaucoma | 70.5 (55.8 - 81.8) | 99.3 (98.8 - 99.6) | 68.9 (54.3 - 80.5) | 99.3 (98.9 - 99.6) | 69.7 (57.5 - 79.4) | 98.6 (98.0 - 99.1) |
|  |  | N/A | 94.0 (91.8 - 95.6) | 98.5 (97.7 - 99.0) | 96.5 (94.7 - 97.7) | 97.3 (96.3 - 98.1) | 95.2 (94.0 - 96.4) | 97.1 (96.2 - 97.7) |
| GPT | 0 (0.0%) | POAG | 98.3 (97.1 - 99.0) | 97.1 (96.0 - 97.9) | 95.9 (94.3 - 97.0) | 98.8 (98.0 - 99.3) | 97.0 (96.2 - 97.8) | 97.6 (96.8 - 98.2) |
|  |  | NTG | 96.1 (91.1 - 98.3) | 99.4 (98.9 - 99.7) | 91.7 (85.8 - 95.3) | 99.7 (99.4 - 99.9) | 93.8 (90.4 - 96.6) | 99.2 (98.7 - 99.5) |
|  |  | PACG | 96.2 (90.6 - 98.5) | 99.3 (98.8 - 99.6) | 87.8 (80.6 - 92.6) | 99.8 (99.5 - 99.9) | 91.8 (87.9 - 95.3) | 99.1 (98.6 - 99.4) |
|  |  | Pseudoexfoliative Glaucoma | 87.6 (79.6 - 92.8) | 99.9 (99.7 - 100.0) | 98.8 (93.7 - 99.8) | 99.4 (98.9 - 99.6) | 92.9 (88.9 - 96.3) | 99.3 (98.9 - 99.6) |
|  |  | Pigmentary Glaucoma | 100.0 (83.9 - 100.0) | 100.0 (99.8 - 100.0) | 100.0 (83.9 - 100.0) | 100.0 (99.8 - 100.0) | 100.0 (100.0 - 100.0) | 100.0 (99.8 - 100.0) |
|  |  | Angle Recession Glaucoma | 75.0 (40.9 - 92.9) | 99.9 (99.7 - 100.0) | 85.7 (48.7 - 97.4) | 99.9 (99.6 - 100.0) | 80.0 (44.4 - 100.0) | 99.8 (99.6 - 99.9) |
|  |  | Uveitic Glaucoma | 81.0 (60.0 - 92.3) | 99.7 (99.3 - 99.9) | 73.9 (53.5 - 87.5) | 99.8 (99.5 - 99.9) | 77.3 (61.1 - 89.8) | 99.5 (99.1 - 99.7) |
|  |  | Steroid-Induced Glaucoma | 93.3 (70.2 - 98.8) | 100.0 (99.8 - 100.0) | 100.0 (78.5 - 100.0) | 99.9 (99.7 - 100.0) | 96.6 (87.0 - 100.0) | 99.9 (99.7 - 100.0) |
|  |  | Neovascular Glaucoma | 100.0 (89.0 - 100.0) | 99.8 (99.5 - 99.9) | 88.6 (74.0 - 95.5) | 100.0 (99.8 - 100.0) | 93.9 (86.8 - 98.6) | 99.8 (99.5 - 99.9) |
|  |  | Mixed Mechanism Glaucoma | 81.8 (70.9 - 89.3) | 99.8 (99.5 - 99.9) | 93.1 (83.6 - 97.3) | 99.4 (98.9 - 99.6) | 87.1 (80.4 - 93.1) | 99.2 (98.7 - 99.5) |
|  |  | Glaucoma following cataract surgery | 85.7 (48.7 - 97.4) | 99.9 (99.7 - 100.0) | 85.7 (48.7 - 97.4) | 99.9 (99.7 - 100.0) | 85.7 (50.0 - 100.0) | 99.9 (99.6 - 100.0) |
|  |  | ICE Syndrome | 80.0 (37.6 - 96.4) | 100.0 (99.8 - 100.0) | 100.0 (51.0 - 100.0) | 99.9 (99.7 - 100.0) | 88.9 (50.0 - 100.0) | 99.9 (99.7 - 100.0) |
|  |  | Sturge-Weber Glaucoma | 100.0 (20.7 - 100.0) | 100.0 (99.8 - 100.0) | 100.0 (20.7 - 100.0) | 100.0 (99.8 - 100.0) | 100.0 (100.0 - 100.0) | 100.0 (99.8 - 100.0) |
|  |  | Other | 90.9 (72.2 - 97.5) | 99.5 (99.1 - 99.7) | 66.7 (48.8 - 80.8) | 99.9 (99.6 - 100.0) | 76.9 (61.9 - 88.5) | 99.4 (98.9 - 99.7) |
|  |  | Congenital Glaucoma | — | 99.9 (99.6 - 100.0) | 0.0 (0.0 - 65.8) | 100.0 (99.8 - 100.0) | — | 99.9 (99.6 - 100.0) |
|  |  | Unspecified Glaucoma | 70.5 (55.8 - 81.8) | 99.2 (98.7 - 99.5) | 66.0 (51.7 - 77.8) | 99.3 (98.9 - 99.6) | 68.1 (55.9 - 77.4) | 98.5 (97.9 - 99.0) |
|  |  | N/A | 92.7 (90.3 - 94.5) | 99.1 (98.5 - 99.5) | 97.9 (96.4 - 98.8) | 96.8 (95.7 - 97.6) | 95.2 (94.0 - 96.4) | 97.1 (96.3 - 97.8) |
| Grok | 0 (0.0%) | POAG | 98.8 (97.7 - 99.3) | 96.3 (95.0 - 97.2) | 94.7 (93.0 - 96.0) | 99.1 (98.4 - 99.5) | 96.7 (95.8 - 97.5) | 97.3 (96.5 - 97.9) |
|  |  | NTG | 86.6 (79.6 - 91.5) | 99.7 (99.4 - 99.9) | 95.7 (90.2 - 98.1) | 99.1 (98.5 - 99.4) | 90.9 (86.6 - 94.4) | 98.9 (98.3 - 99.3) |
|  |  | PACG | 91.4 (84.5 - 95.4) | 99.8 (99.5 - 99.9) | 96.0 (90.2 - 98.4) | 99.5 (99.1 - 99.7) | 93.7 (90.0 - 96.8) | 99.3 (98.9 - 99.6) |
|  |  | Pseudoexfoliative Glaucoma | 88.7 (80.8 - 93.5) | 99.9 (99.7 - 100.0) | 98.9 (93.8 - 99.8) | 99.4 (99.0 - 99.7) | 93.5 (89.3 - 96.7) | 99.4 (98.9 - 99.7) |
|  |  | Pigmentary Glaucoma | 90.0 (69.9 - 97.2) | 99.9 (99.7 - 100.0) | 94.7 (75.4 - 99.1) | 99.9 (99.6 - 100.0) | 92.3 (81.2 - 100.0) | 99.8 (99.6 - 99.9) |
|  |  | Angle Recession Glaucoma | 87.5 (52.9 - 97.8) | 100.0 (99.8 - 100.0) | 100.0 (64.6 - 100.0) | 99.9 (99.7 - 100.0) | 93.3 (72.7 - 100.0) | 99.9 (99.7 - 100.0) |
|  |  | Uveitic Glaucoma | 95.2 (77.3 - 99.2) | 99.7 (99.4 - 99.9) | 80.0 (60.9 - 91.1) | 99.9 (99.7 - 100.0) | 87.0 (75.7 - 96.0) | 99.7 (99.3 - 99.9) |
|  |  | Steroid-Induced Glaucoma | 53.3 (30.1 - 75.2) | 100.0 (99.8 - 100.0) | 100.0 (67.6 - 100.0) | 99.6 (99.3 - 99.8) | 69.6 (40.0 - 88.0) | 99.6 (99.3 - 99.8) |
|  |  | Neovascular Glaucoma | 100.0 (89.0 - 100.0) | 99.9 (99.7 - 100.0) | 96.9 (84.3 - 99.4) | 100.0 (99.8 - 100.0) | 98.4 (94.5 - 100.0) | 99.9 (99.7 - 100.0) |
|  |  | Mixed Mechanism Glaucoma | 83.3 (72.6 - 90.4) | 99.9 (99.7 - 100.0) | 98.2 (90.6 - 99.7) | 99.4 (99.0 - 99.7) | 90.2 (84.0 - 95.1) | 99.4 (98.9 - 99.7) |
|  |  | Glaucoma following cataract surgery | 100.0 (64.6 - 100.0) | 100.0 (99.8 - 100.0) | 100.0 (64.6 - 100.0) | 100.0 (99.8 - 100.0) | 100.0 (100.0 - 100.0) | 100.0 (99.8 - 100.0) |
|  |  | ICE Syndrome | 60.0 (23.1 - 88.2) | 100.0 (99.8 - 100.0) | 100.0 (43.9 - 100.0) | 99.9 (99.6 - 100.0) | 75.0 (0.0 - 100.0) | 99.9 (99.6 - 100.0) |
|  |  | Sturge-Weber Glaucoma | 100.0 (20.7 - 100.0) | 100.0 (99.8 - 100.0) | 100.0 (20.7 - 100.0) | 100.0 (99.8 - 100.0) | 100.0 (100.0 - 100.0) | 100.0 (99.8 - 100.0) |
|  |  | Other | 72.7 (51.8 - 86.8) | 99.4 (99.0 - 99.7) | 59.3 (40.7 - 75.5) | 99.7 (99.3 - 99.9) | 65.3 (46.2 - 79.2) | 99.1 (98.6 - 99.5) |
|  |  | Congenital Glaucoma | — | 99.8 (99.6 - 99.9) | 0.0 (0.0 - 56.1) | 100.0 (99.8 - 100.0) | — | 99.8 (99.6 - 99.9) |
|  |  | Unspecified Glaucoma | 61.4 (46.6 - 74.3) | 99.7 (99.3 - 99.9) | 81.8 (65.6 - 91.4) | 99.1 (98.6 - 99.5) | 70.1 (56.7 - 80.0) | 98.8 (98.3 - 99.2) |
|  |  | N/A | 95.9 (94.1 - 97.2) | 97.3 (96.3 - 98.0) | 94.1 (92.0 - 95.7) | 98.2 (97.3 - 98.7) | 95.0 (93.8 - 96.2) | 96.9 (96.0 - 97.6) |
| Qwen | 52 (5.2%) | POAG | 99.5 (98.7 - 99.8) | 95.4 (93.9 - 96.4) | 93.9 (92.0 - 95.3) | 99.6 (99.0 - 99.9) | 96.6 (95.7 - 97.4) | 97.1 (96.2 - 97.7) |
|  |  | NTG | 95.9 (90.8 - 98.3) | 99.5 (99.1 - 99.8) | 93.7 (88.0 - 96.7) | 99.7 (99.3 - 99.9) | 94.8 (91.7 - 97.5) | 99.3 (98.8 - 99.6) |
|  |  | PACG | 98.0 (92.9 - 99.4) | 99.2 (98.7 - 99.5) | 87.4 (79.9 - 92.3) | 99.9 (99.6 - 100.0) | 92.4 (88.6 - 95.6) | 99.1 (98.6 - 99.5) |
|  |  | Pseudoexfoliative Glaucoma | 88.6 (80.3 - 93.7) | 100.0 (99.8 - 100.0) | 100.0 (95.3 - 100.0) | 99.4 (99.0 - 99.7) | 94.0 (89.7 - 97.3) | 99.5 (99.0 - 99.7) |
|  |  | Pigmentary Glaucoma | 100.0 (83.9 - 100.0) | 99.9 (99.7 - 100.0) | 95.2 (77.3 - 99.2) | 100.0 (99.8 - 100.0) | 97.6 (91.7 - 100.0) | 99.9 (99.7 - 100.0) |
|  |  | Angle Recession Glaucoma | 100.0 (64.6 - 100.0) | 99.9 (99.7 - 100.0) | 87.5 (52.9 - 97.8) | 100.0 (99.8 - 100.0) | 93.3 (72.7 - 100.0) | 99.9 (99.7 - 100.0) |
|  |  | Uveitic Glaucoma | 94.4 (74.2 - 99.0) | 99.7 (99.4 - 99.9) | 77.3 (56.6 - 89.9) | 99.9 (99.7 - 100.0) | 85.0 (70.8 - 95.0) | 99.7 (99.3 - 99.9) |
|  |  | Steroid-Induced Glaucoma | 86.7 (62.1 - 96.3) | 100.0 (99.8 - 100.0) | 100.0 (77.2 - 100.0) | 99.9 (99.6 - 100.0) | 92.9 (80.0 - 100.0) | 99.9 (99.6 - 100.0) |
|  |  | Neovascular Glaucoma | 100.0 (87.5 - 100.0) | 99.9 (99.7 - 100.0) | 96.4 (82.3 - 99.4) | 100.0 (99.8 - 100.0) | 98.2 (93.3 - 100.0) | 99.9 (99.7 - 100.0) |
|  |  | Mixed Mechanism Glaucoma | 85.0 (73.9 - 91.9) | 100.0 (99.8 - 100.0) | 100.0 (93.0 - 100.0) | 99.5 (99.1 - 99.7) | 91.9 (85.5 - 96.6) | 99.5 (99.1 - 99.7) |
|  |  | Glaucoma following cataract surgery | 100.0 (61.0 - 100.0) | 100.0 (99.8 - 100.0) | 100.0 (61.0 - 100.0) | 100.0 (99.8 - 100.0) | 100.0 (100.0 - 100.0) | 100.0 (99.8 - 100.0) |
|  |  | ICE Syndrome | 75.0 (30.1 - 95.4) | 100.0 (99.8 - 100.0) | 100.0 (43.9 - 100.0) | 99.9 (99.7 - 100.0) | 85.7 (0.0 - 100.0) | 99.9 (99.7 - 100.0) |
|  |  | Sturge-Weber Glaucoma | 0.0 (0.0 - 79.3) | 100.0 (99.8 - 100.0) | — | 99.9 (99.7 - 100.0) | 0.0 (0.0 - 0.0) | 99.9 (99.7 - 100.0) |
|  |  | Other | 80.0 (58.4 - 91.9) | 99.5 (99.1 - 99.7) | 64.0 (44.5 - 79.8) | 99.8 (99.4 - 99.9) | 71.1 (54.5 - 85.0) | 99.3 (98.8 - 99.6) |
|  |  | Congenital Glaucoma | — | 99.8 (99.5 - 99.9) | 0.0 (0.0 - 49.0) | 100.0 (99.8 - 100.0) | — | 99.8 (99.5 - 99.9) |
|  |  | Unspecified Glaucoma | 70.3 (54.2 - 82.5) | 99.8 (99.5 - 99.9) | 89.7 (73.6 - 96.4) | 99.4 (98.9 - 99.7) | 78.8 (65.6 - 88.6) | 99.3 (98.8 - 99.6) |
|  |  | N/A | 90.4 (87.7 - 92.5) | 99.4 (98.8 - 99.7) | 98.5 (97.0 - 99.2) | 95.9 (94.8 - 96.9) | 94.3 (92.8 - 95.6) | 96.7 (95.7 - 97.4) |

95% confidence intervals are reported in parentheses. — = undefined: the class was never predicted (PPV) or was absent from the test set (sensitivity and F1).
NPV = negative predictive value; NTG = Normal Tension Glaucoma; PACG = Primary Angle-Closure Glaucoma; PPV = positive predictive value; POAG = Primary Open-Angle Glaucoma.

**Table 4.** Diagnostic performance of ICD-10-CM codes for glaucoma type classification.

| **Glaucoma Type** | **Sensitivity<br>(%)** | **Specificity<br>(%)** | **PPV<br>(%)** | **NPV<br>(%)** | **F1-Score<br>(%)** | **Accuracy<br>(%)** |
|---|---|---|---|---|---|---|
| POAG | 90.5<br>(88.3 - 92.3) | 89.9<br>(88.0 - 91.5) | 85.8<br>(83.3 - 88.0) | 93.3<br>(91.7 - 94.6) | 88.1<br>(86.4 - 89.6) | 90.1<br>(88.7 - 91.4) |
| NTG | 89.0<br>(82.4 - 93.3) | 99.0<br>(98.5 - 99.4) | 86.3<br>(79.3 - 91.1) | 99.2<br>(98.7 - 99.6) | 87.6<br>(83.0 - 91.1) | 98.4<br>(97.7 - 98.9) |
| PACG | 84.8<br>(76.7 - 90.4) | 98.1<br>(97.4 - 98.6) | 71.2<br>(62.7 - 78.4) | 99.1<br>(98.6 - 99.5) | 77.4<br>(71.6 - 82.3) | 97.4<br>(96.6 - 98.0) |
| Pseudoexfoliative Glaucoma | 69.1<br>(59.3 - 77.4) | 99.7<br>(99.3 - 99.9) | 91.8<br>(83.2 - 96.2) | 98.4<br>(97.8 - 98.9) | 78.8<br>(72.1 - 84.3) | 98.2<br>(97.5 - 98.7) |
| Pigmentary Glaucoma | 70.0<br>(48.1 - 85.5) | 99.9<br>(99.6 - 100.0) | 87.5<br>(64.0 - 96.5) | 99.7<br>(99.3 - 99.9) | 77.8<br>(61.9 - 88.3) | 99.6<br>(99.2 - 99.8) |
| Angle Recession Glaucoma | 75.0<br>(40.9 - 92.9) | 99.9<br>(99.6 - 100.0) | 66.7<br>(35.4 - 87.9) | 99.9<br>(99.6 - 100.0) | 70.6<br>(46.9 - 86.7) | 99.8<br>(99.4 - 99.9) |
| Uveitic Glaucoma | 71.4<br>(50.0 - 86.2) | 99.4<br>(99.0 - 99.7) | 57.7<br>(39.0 - 74.5) | 99.7<br>(99.3 - 99.9) | 63.8<br>(49.5 - 76.0) | 99.1<br>(98.6 - 99.5) |

95% confidence intervals are reported in parentheses. Absent documentation in either gold standard or ICD code was considered negative for the specific condition.
ICD-10-CM = International Classification of Diseases, 10th Revision, Clinical Modification; NPV = negative predictive value; NTG = Normal Tension Glaucoma; PACG = Primary Angle-Closure Glaucoma; PPV = positive predictive value; POAG = Primary Open-Angle Glaucoma.

For glaucoma severity staging, overall accuracy ranged from 94.0% (95% CI: 92.9% - 95.0%) to 95.2% (95% CI: 94.1% - 96.0%). Overall accuracies were 95.0% (95% CI: 93.9% - 95.9%), 94.8% (95% CI: 93.7% - 95.7%), 94.5% (95% CI: 93.4% - 95.4%), 94.0% (95% CI: 92.9% - 95.0%), and 95.2% (95% CI: 94.1% - 96.0%) for Claude, DeepSeek, GPT, Grok and Qwen, respectively (**Table 5**). F1 scores ranged from 89.7% to 96.3%. Failure rates ranged from 0% to 2.2% across models, with Qwen exhibiting the highest failure rate. Severity classification errors involved confusion between adjacent stages, such as mild versus moderate or moderate versus severe disease, and misclassification between defined severity stages and the ’Other’ category (Figure 3).

**Figure 3.**
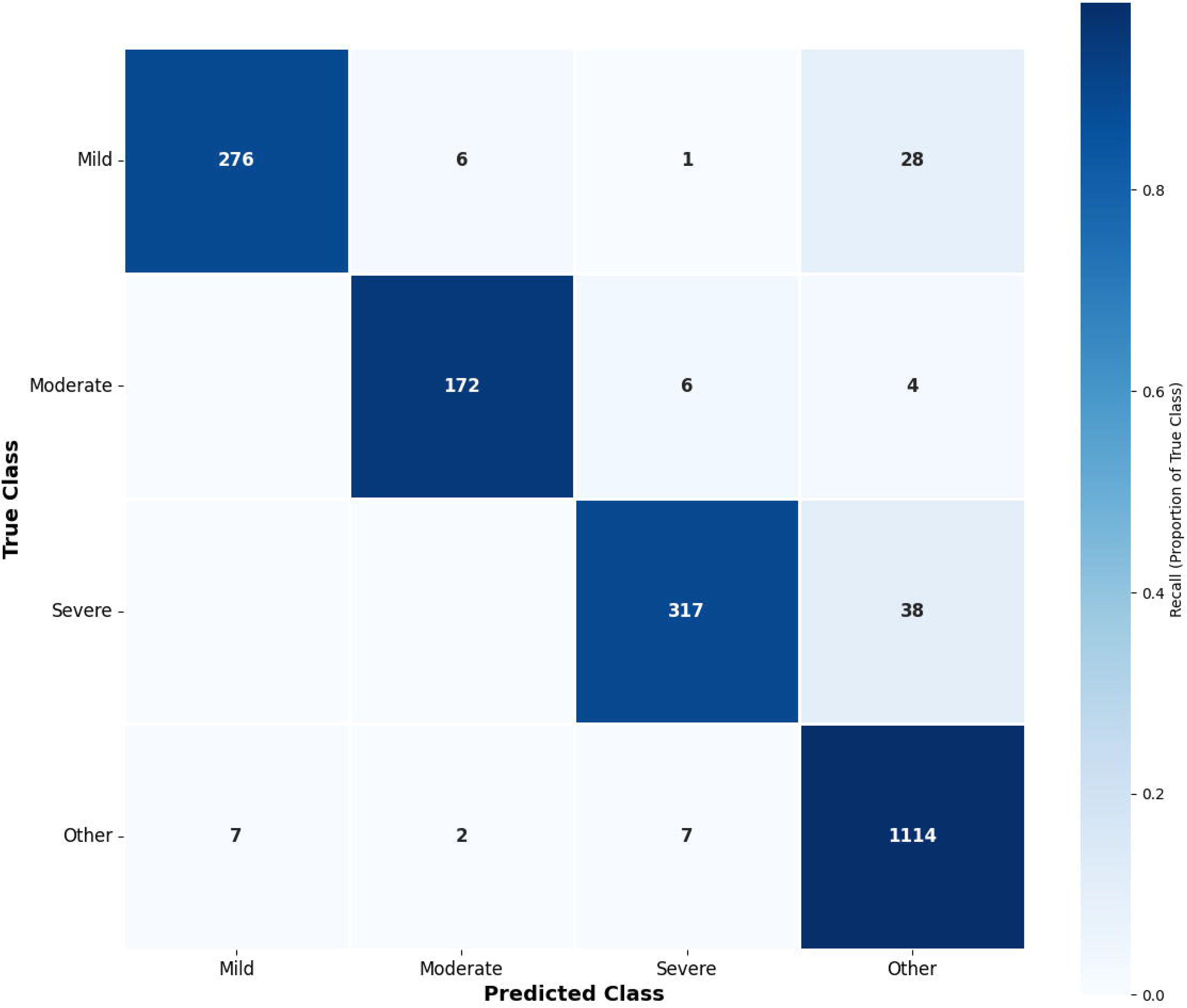
Confusion matrix for glaucoma severity classification using output from Claude. A total of 6 eyes from 3 notes with failed outputs were excluded.

**Table 5.**
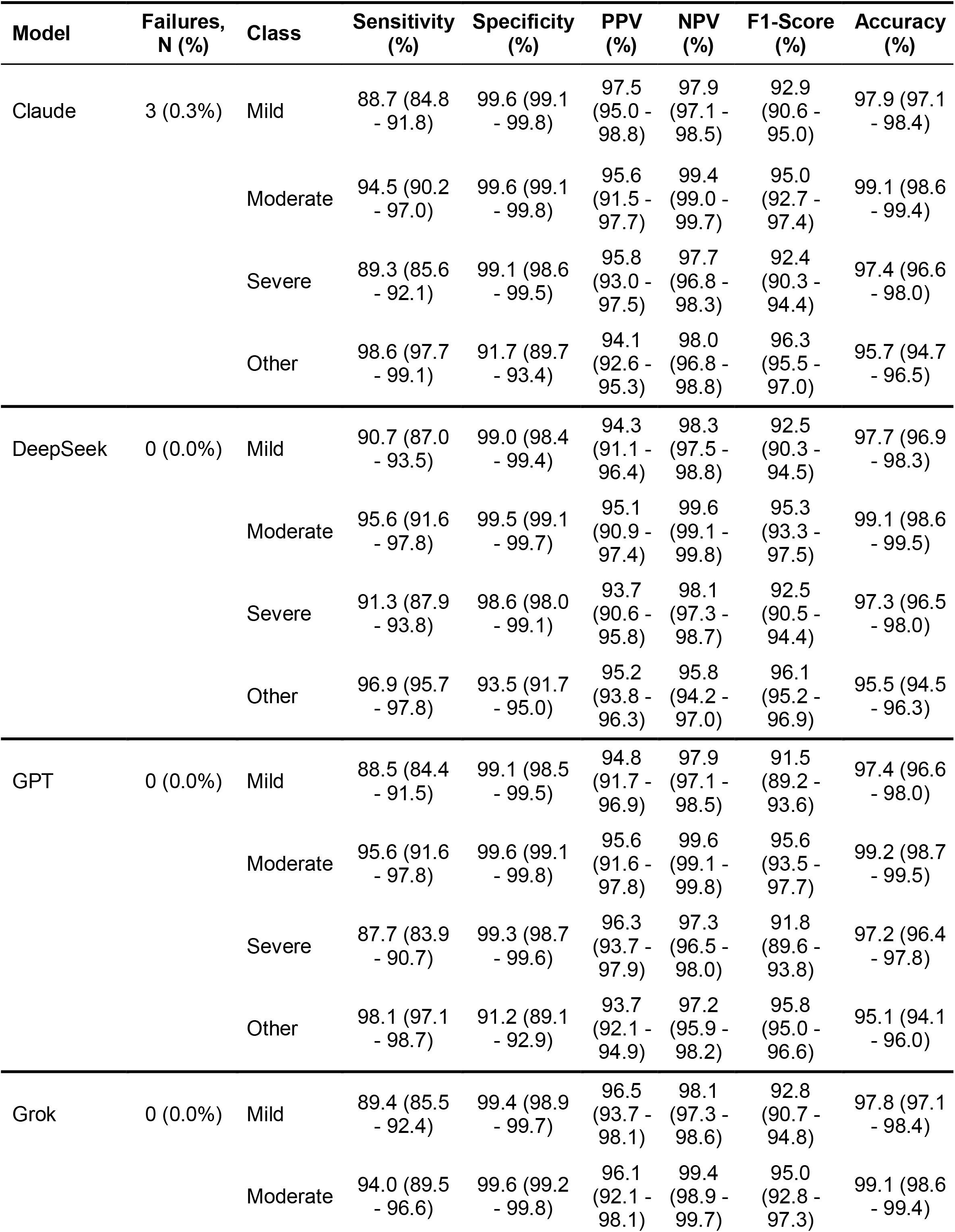
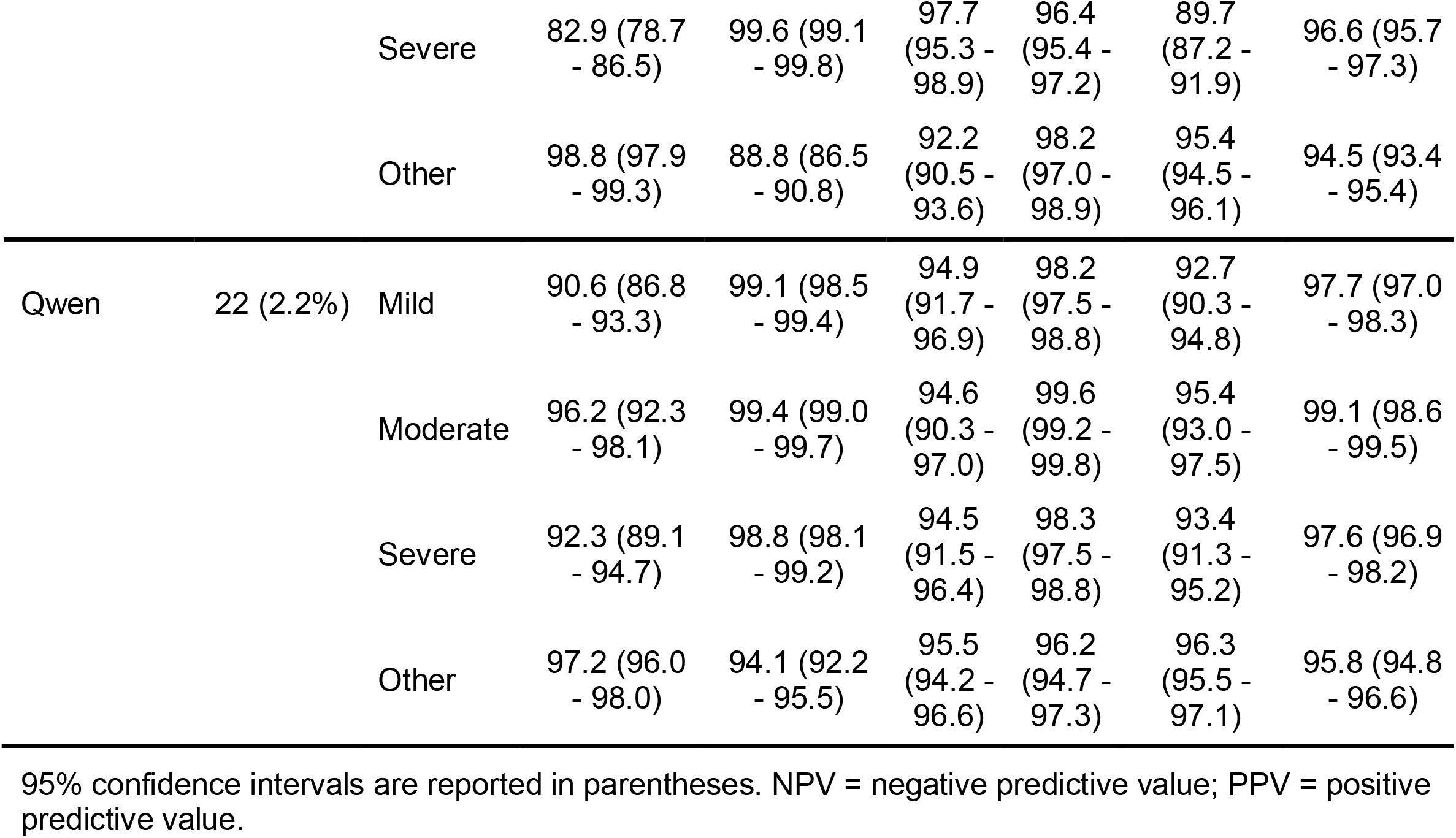
Class-specific performance of large language models for glaucoma severity classification.

| **Model** | **Failures, N (%)** | **Class** | **Sensitivity (%)** | **Specificity (%)** | **PPV (%)** | **NPV (%)** | **F1-Score (%)** | **Accuracy (%)** |
|---|---|---|---|---|---|---|---|---|
| Claude | 3 (0.3%) | Mild | 88.7 (84.8 - 91.8) | 99.6 (99.1 - 99.8) | 97.5 (95.0 - 98.8) | 97.9 (97.1 - 98.5) | 92.9 (90.6 - 95.0) | 97.9 (97.1 - 98.4) |
|  |  | Moderate | 94.5 (90.2 - 97.0) | 99.6 (99.1 - 99.8) | 95.6 (91.5 - 97.7) | 99.4 (99.0 - 99.7) | 95.0 (92.7 - 97.4) | 99.1 (98.6 - 99.4) |
|  |  | Severe | 89.3 (85.6 - 92.1) | 99.1 (98.6 - 99.5) | 95.8 (93.0 - 97.5) | 97.7 (96.8 - 98.3) | 92.4 (90.3 - 94.4) | 97.4 (96.6 - 98.0) |
|  |  | Other | 98.6 (97.7 - 99.1) | 91.7 (89.7 - 93.4) | 94.1 (92.6 - 95.3) | 98.0 (96.8 - 98.8) | 96.3 (95.5 - 97.0) | 95.7 (94.7 - 96.5) |
| DeepSeek | 0 (0.0%) | Mild | 90.7 (87.0 - 93.5) | 99.0 (98.4 - 99.4) | 94.3 (91.1 - 96.4) | 98.3 (97.5 - 98.8) | 92.5 (90.3 - 94.5) | 97.7 (96.9 - 98.3) |
|  |  | Moderate | 95.6 (91.6 - 97.8) | 99.5 (99.1 - 99.7) | 95.1 (90.9 - 97.4) | 99.6 (99.1 - 99.8) | 95.3 (93.3 - 97.5) | 99.1 (98.6 - 99.5) |
|  |  | Severe | 91.3 (87.9 - 93.8) | 98.6 (98.0 - 99.1) | 93.7 (90.6 - 95.8) | 98.1 (97.3 - 98.7) | 92.5 (90.5 - 94.4) | 97.3 (96.5 - 98.0) |
|  |  | Other | 96.9 (95.7 - 97.8) | 93.5 (91.7 - 95.0) | 95.2 (93.8 - 96.3) | 95.8 (94.2 - 97.0) | 96.1 (95.2 - 96.9) | 95.5 (94.5 - 96.3) |
| GPT | 0 (0.0%) | Mild | 88.5 (84.4 - 91.5) | 99.1 (98.5 - 99.5) | 94.8 (91.7 - 96.9) | 97.9 (97.1 - 98.5) | 91.5 (89.2 - 93.6) | 97.4 (96.6 - 98.0) |
|  |  | Moderate | 95.6 (91.6 - 97.8) | 99.6 (99.1 - 99.8) | 95.6 (91.6 - 97.8) | 99.6 (99.1 - 99.8) | 95.6 (93.5 - 97.7) | 99.2 (98.7 - 99.5) |
|  |  | Severe | 87.7 (83.9 - 90.7) | 99.3 (98.7 - 99.6) | 96.3 (93.7 - 97.9) | 97.3 (96.5 - 98.0) | 91.8 (89.6 - 93.8) | 97.2 (96.4 - 97.8) |
|  |  | Other | 98.1 (97.1 - 98.7) | 91.2 (89.1 - 92.9) | 93.7 (92.1 - 94.9) | 97.2 (95.9 - 98.2) | 95.8 (95.0 - 96.6) | 95.1 (94.1 - 96.0) |
| Grok | 0 (0.0%) | Mild | 89.4 (85.5 - 92.4) | 99.4 (98.9 - 99.7) | 96.5 (93.7 - 98.1) | 98.1 (97.3 - 98.6) | 92.8 (90.7 - 94.8) | 97.8 (97.1 - 98.4) |
|  |  | Moderate | 94.0 (89.5 - 96.6) | 99.6 (99.2 - 99.8) | 96.1 (92.1 - 98.1) | 99.4 (98.9 - 99.7) | 95.0 (92.8 - 97.3) | 99.1 (98.6 - 99.4) |
|  |  | Severe | 82.9 (78.7 - 86.5) | 99.6 (99.1 - 99.8) | 97.7 (95.3 - 98.9) | 96.4 (95.4 - 97.2) | 89.7 (87.2 - 91.9) | 96.6 (95.7 - 97.3) |
|  |  | Other | 98.8 (97.9 - 99.3) | 88.8 (86.5 - 90.8) | 92.2 (90.5 - 93.6) | 98.2 (97.0 - 98.9) | 95.4 (94.5 - 96.1) | 94.5 (93.4 - 95.4) |
| Qwen | 22 (2.2%) | Mild | 90.6 (86.8 - 93.3) | 99.1 (98.5 - 99.4) | 94.9 (91.7 - 96.9) | 98.2 (97.5 - 98.8) | 92.7 (90.3 - 94.8) | 97.7 (97.0 - 98.3) |
|  |  | Moderate | 96.2 (92.3 - 98.1) | 99.4 (99.0 - 99.7) | 94.6 (90.3 - 97.0) | 99.6 (99.2 - 99.8) | 95.4 (93.0 - 97.5) | 99.1 (98.6 - 99.5) |
|  |  | Severe | 92.3 (89.1 - 94.7) | 98.8 (98.1 - 99.2) | 94.5 (91.5 - 96.4) | 98.3 (97.5 - 98.8) | 93.4 (91.3 - 95.2) | 97.6 (96.9 - 98.2) |
|  |  | Other | 97.2 (96.0 - 98.0) | 94.1 (92.2 - 95.5) | 95.5 (94.2 - 96.6) | 96.2 (94.7 - 97.3) | 96.3 (95.5 - 97.1) | 95.8 (94.8 - 96.6) |

95% confidence intervals are reported in parentheses. NPV = negative predictive value; PPV = positive predictive value.

For severity classification, ICD codes showed weak overall agreement with expert grading, with an overall accuracy of 69.2% (95% CI 67.1–71.2%). ICD code performance varied across severity categories, as shown in **Table 6**. For mild disease, sensitivity was 70.8% and specificity was 91.6%, with an accuracy of 88.4% (95% CI 86.9–89.7%) and F1 score of 65.7% (95% CI 62.0% - 69.2%). For moderate glaucoma, ICD codes demonstrated lower discriminative performance, with an F1-score of 47.2% (95% CI 43.0% - 51.4%) driven by a low positive predictive value (35.9%). Severe disease classification had a sensitivity of 71.4% and specificity of 95.1%, resulting in an accuracy of 90.8% (95% CI 89.5–92.0%) and an F1-score of 73.7% (95% CI 70.3–76.8%). For cases coded as ‘Other’, ICD codes performed poorly, with sensitivity of only 68.1%, corresponding to an accuracy of 73.3% (95% CI 71.4% - 75.2%) and an F1-score of 74.5% (95% CI 72.6% - 76.3%).

**Table 6.** Diagnostic performance of ICD-10-CM codes for glaucoma severity classification.

| Severity | Sensitivity (%) | Specificity (%) | PPV (%) | NPV (%) | F1-Score (%) | Accuracy (%) |
|---|---|---|---|---|---|---|
| Mild | 70.8 (65.6 - 75.6) | 91.6 (90.2 - 92.9) | 61.2 (56.1 - 66.1) | 94.4 (93.2 - 95.4) | 65.7 (62.0 - 69.2) | 88.4 (86.9 - 89.7) |
| Moderate | 68.7 (61.6 - 75.0) | 87.6 (86.0 - 89.1) | 35.9 (31.1 - 41.1) | 96.5 (95.5 - 97.3) | 47.2 (43.0 - 51.4) | 85.9 (84.3 - 87.4) |
| Severe | 71.4 (66.5 - 75.9) | 95.1 (93.9 - 96.0) | 76.1 (71.3 - 80.4) | 93.8 (92.6 - 94.9) | 73.7 (70.3 - 76.8) | 90.8 (89.5 - 92.0) |
| Other | 68.1 (65.4 - 70.8) | 80.3 (77.5 - 82.8) | 82.1 (79.6 - 84.4) | 65.4 (62.5 - 68.3) | 74.5 (72.6 - 76.3) | 73.3 (71.4 - 75.2) |

95% confidence intervals are reported in parentheses. Absent documentation in either gold standard or ICD code was considered negative for the specific condition. ICD-10-CM = International Classification of Diseases, 10th Revision, Clinical Modification; NPV = negative predictive value; PPV = positive predictive value.

When assessing NLP performance (Table 7), BioClinicalBERT underperformed each LLM across all tasks. For diagnosis, accuracy was 68.3% (95% CI: 66.4-70.3%; macro-F1 43.8%; κ 0.31). For type, accuracy was 52.2% (95% CI: 49.9-54.4%; macro-F1 9.0%; κ 0.27). For severity, accuracy was 52.5% (95% CI: 50.4-54.8%; macro-F1 34.7%; κ 0.18), notably worse than ICD-10 codes (69.2%) as well. F1 scores for mild, moderate, and severe disease ranged from 21.6% to 23.6%, with predictions concentrated in the majority “other” class. Supplementary Table S1 provides performance metrics per class for the three tasks.

**Table 7.** Performance of fine-tuned BioClinicalBERT for glaucoma diagnosis, type, and severity classification.

| **Task** | **Accuracy (%)** | **Balanced accuracy (%)** | **Macro F1 (%)** | **Weighted F1 (%)** | **Cohen’s κ** |
|---|---|---|---|---|---|
| Diagnosis | 68.3<br>(66.4 - 70.3) | 46.7<br>(44.9 - 48.4) | 43.8<br>(42.1 - 45.4) | 67.0<br>(64.8 - 69.0) | 0.31<br>(0.27 - 0.35) |
| Type | 52.2<br>(49.9 - 54.4) | 10.1<br>(9.6 - 11.2) | 9.0<br>(8.5 - 10.1) | 45.3<br>(43.0 - 47.6) | 0.27<br>(0.24 - 0.30) |
| Severity | 52.5<br>(50.4 - 54.8) | 34.7<br>(32.5 - 36.8) | 34.7<br>(32.4 - 37.0) | 50.6<br>(48.2 - 53.0) | 0.18<br>(0.15 - 0.21) |

95% confidence intervals are reported in parentheses. Balanced accuracy is the mean class-specific sensitivity; macro F1 is the unweighted mean of the class-specific F1 scores; weighted F1 weights each class-specific F1 by the number of eyes in that class. Balanced accuracy and macro F1 were averaged over classes present in the test set.

## DISCUSSION

In this study, secure cloud-based LLMs performed with high accuracy in extracting glaucoma diagnosis, type, and severity information from free-text EHR clinical notes. Across all models, performance was consistently high for all tasks. For glaucoma diagnosis, overall accuracy ranged from 94.4% to 97.5%, while for type classification, accuracies ranged from 94.0% to 97.1%. Severity classification accuracies ranged from 94.0% to 95.2%. By converting narrative documentation into structured variables at scale, LLMs address known limitations of relying solely on billing codes or sparse structured EHR fields for disease phenotype construction and enable downstream analytics that require precise assessments of disease presence, type, and severity level. Importantly, these findings demonstrate that detailed glaucoma phenotypes can be extracted directly from routine clinical documentation without ophthalmology-specific model development or specialty-specific pretraining. By enabling scalable extraction of clinically meaningful disease phenotypes, LLM-based workflows operating within a HIPAA-compliant infrastructure may substantially improve the quality of glaucoma cohort definitions for large-scale research applications.

Accurate characterization of glaucoma diagnosis, type, and severity is essential to large-scale disease phenotyping. Misclassification at any of these levels introduces bias into cohort definitions, distorts prevalence estimates, and impairs the validity of stage-stratified analyses central to glaucoma research.^5, 6^ Severity staging informs the intensity of therapeutic intervention as well as the frequency of follow-up, and is frequently employed as an eligibility criterion in clinical trials and longitudinal cohort studies. Similarly, accurate type identification may be essential for genetic association studies and multimodal modeling efforts.^3, 25–27^ Our findings suggest that LLMs can successfully recover this information directly from free-text documentation with performance approaching expert clinician adjudication. Importantly, unlike traditional rule-based NLP pipelines, LLMs are capable of contextual interpretation of complex clinical narratives, allowing them to synthesize dispersed, potentially conflicting information embedded throughout the note to produce the most likely output.^13^ Of note, we considered allowing the graders to infer severity by analyzing perimetric findings included in the clinical note. However, we decided to not pursue this for three reasons: 1) perimetric findings can be confounded by other ocular diseases, which may lead the clinician to intentionally refrain from explicitly stating the severity; 2) given the goal of applications in research, requirement of an explicit descriptor by the clinician would ensure the most conservative output; 3) we were concerned about potential hallucinations by the models. With these restrictions, nearly two-fifths of glaucomatous eyes still had unspecified/indeterminate severity per adjudicated labels.

Model performance was consistently high across all five LLMs. Claude achieved the highest overall accuracy for glaucoma diagnosis (97.5%) and type classification (97.1%), while Qwen performed best in severity staging (95.2%). Similar trends were noticeable with F1 scores, with Claude achieving the highest F1 scores for common glaucoma types, such as POAG (98.9%) and NTG (96.1%), though performance was more variable for categories with limited case representation, such as uveitic glaucoma (73.2%) given lower prevalence. Grok demonstrated the lowest overall accuracy, driven in part by reduced PPV for the "Neither" class, suggesting a modest tendency toward over-labeling. DeepSeek and GPT performed comparably across all tasks. These findings suggest task-specific differences in observed performance, although formal statistical comparisons between models were not performed and failure rates differed between models. Notably, failure rates were higher with Claude and Qwen; we hypothesize that this pattern may reflect differences in model architecture, with Claude and Qwen potentially employing more extensive deductive reasoning that increased token use and led to output failures when token limits were reached.

A major finding of this study was the substantial performance gap between LLM-based phenotyping and ICD-based classification, particularly for glaucoma severity staging. ICD-10-CM codes demonstrated relatively poor performance for severity classification, with an overall accuracy of only 69.2%, largely driven by the frequent use of unspecified staging codes. The low sensitivities of mild, moderate, and severe stages reflect the suboptimal use of precise staging in ICD codes (Table 6). In contrast, all five LLMs achieved approximately 95% accuracies for severity staging with high sensitivities and specificities (**Table 5**). Similar differences were observed for glaucoma type classification, in which ICD-based approaches demonstrated lower sensitivity for less prevalent glaucoma phenotypes, including pseudoexfoliative glaucoma (sensitivity 69.1%, F1-score 78.8%), pigmentary glaucoma (sensitivity 70.0%, F1-score 77.8%), angle recession glaucoma (sensitivity 75.0%, F1-score 70.6%), and uveitic glaucoma (sensitivity 71.4%, F1-score 63.8%). These findings are consistent with prior literature demonstrating important limitations of ICD-based glaucoma phenotyping.^6, 7, 28^ In routine clinical practice, ICD coding is frequently influenced not only by diagnostic precision but also by administrative and billing considerations, including optimization for insurance authorization and reimbursement. As a result, clinicians may preferentially select broader or nonspecific billing codes even when more clinically precise phenotypic information is available and documented within the note. LLMs can overcome these limitations by extracting clinically meaningful information from narrative documentation. Accurate severity phenotyping is highly relevant for glaucoma research as disease stage often determines eligibility criteria, treatment stratification, progression analyses, and outcome interpretation in both epidemiologic and interventional studies.^3, 6^

Representative examples of misclassifications are provided as **Supplementary Table S2**. Misclassifications tended to cluster around a specific and understandable challenge – navigating the distinction between genuine clinical uncertainty and a clinician’s working estimate. In glaucoma type classification, errors arose when clinicians documented two competing diagnoses (e.g., “POAG/NTG”) in which the model’s rule-based tiebreaker, while consistent, might not have captured the clinician’s intention. For severity classification, errors largely stemmed from notes where a formal indeterminate label co-existed with a parenthetical directional estimate, such as “stage indeterminant (likely mild OD, moderate OS)”. Similarly, with glaucoma diagnosis, misclassifications centered on probabilistic language such as “likely.” Most misclassifications were reflective of the challenge in how classification rules handle possible ambiguities in clinical assessments.

Our study also differs from prior ophthalmic NLP literature in several important ways. Prior NLP studies in glaucoma have demonstrated the value of text-based extraction for medications,^8, 10^ binary diagnosis,^11, 29, 30^ and enhanced phenotyping through multi-source EHR integration.^12^ These approaches have largely relied on task-specific extraction pipelines or conventional machine learning frameworks requiring predefined variables and extensive preprocessing. In this work, BioClinicalBERT reached only 52-68% accuracy across tasks despite long-document encoding and class-weighted training. With limited annotated data, it failed to learn rare glaucoma types or distinguish severity stages, whereas all five LLMs achieved an accuracy of 94% or better through prompting alone. Recent ophthalmology domain-specific encoders such as OphthaBERT have demonstrated that glaucoma diagnosis and type classification can be successfully extracted from narrative documentation,^11^ but it required large-scale specialty pretraining and task-specific fine-tuning. We extend this work by showing that general-purpose LLMs, without domain pretraining or model fine-tuning, can accurately recover not only glaucoma diagnosis and type at a more granular level, but also eye-level disease severity. Given these models operated within a secure HIPAA-compliant cloud environment, our results support the feasibility of deploying these methods for large-scale clinical research involving PHI.

Cloud-based LLMs carry both time and monetary costs. In this study, processing the test set took approximately 20-45 minutes per model for glaucoma diagnosis and 45-90 minutes per model for glaucoma type and severity. Usage was not tracked per model, but total usage across all five models and all three tasks during the 1-month study period was approximately $1,400, a mean of roughly $93 per model per task. Because each note is processed independently, requests can be run in parallel. Scaling to larger datasets is therefore limited mainly by deployment throughput (e.g., API rate limits) and cost rather than by processing time. These estimates reflect vendor pricing and model versions at the time of this study, both which may change.

Several limitations should be considered when interpreting these findings. First, this study was conducted at a single academic institution, and prompt development was performed using clinical notes from the same institution, potentially limiting generalizability to other healthcare systems. Differences in clinical documentation styles, terminology, and EHR workflows may affect extraction performance. Although our prompts contained minimal institution-specific components, external validation using independently collected clinical notes from other institutions is necessary to establish the generalizability of this approach before broader implementation. Second, our cohort included only patient encounters with glaucoma specialists associated with glaucoma-related ICD codes, potentially excluding patients with glaucoma who were not appropriately coded. Visits with non-glaucoma specialists (e.g., comprehensive ophthalmologists or optometrists) were also intentionally excluded in this work, primarily due to concerns of suboptimal documentation regarding type and severity. Therefore, this work currently supports the use of LLMs for improving the accuracy of glaucoma phenotyping among patients cared for by glaucoma specialists. Further validation in broader clinical populations would be needed to establish the performance and generalizability of this approach across diverse practice settings. Access to enterprise-grade secure AI infrastructure currently remains limited, but the availability of HIPAA-compliant cloud-based AI platforms is rapidly expanding and may improve accessibility over time. Additionally, although performance was evaluated at the eye level, within-patient correlation between fellow eyes was not accounted for when estimating CIs. Reported CIs may underestimate statistical uncertainty, although the point estimates of performance remain unaffected. Finally, some rare glaucoma phenotypes contained relatively small sample sizes, limiting precision of class-specific performance estimates. Future studies could include external validation and explorations of multimodal frameworks integrating clinical text with ophthalmic testing data.^26, 27, 31, 32^

In summary, secure cloud-based LLMs consistently demonstrated high accuracy in comprehensively describing glaucoma phenotypes in terms of diagnosis, type, and severity using free-text clinical notes, with performance approaching expert clinician adjudication. Models outperformed traditional ICD-based phenotyping approaches. These findings indicate that modern LLMs can extract multidimensional glaucoma phenotypes from routine clinical notes without specialized training, enabling efficient conversion of unstructured ophthalmology records into structured data for large-scale research.

## Supporting information

Supplemental Material: Prompts

## Data Availability

Data (except raw clinical notes) will be available upon reasonable request

## Acknowledgements

None.

## Abbreviations

AI: Artificial Intelligence
API: Application Programming Interface
HER: Electronic Health Record
LLM: Large Language Model
NLP: Natural language processing
PHI: Protected Health Information
SLM: Small Language Model

